# Predictors of 30-Day Readmission & Reoperations Post Strictureplasty in Crohn’s Disease: A NSQIP Retrospective Review and Analysis

**DOI:** 10.1101/2022.09.16.22279987

**Authors:** Apar S. Patel, Emma K. Satchel, Swadha Guru, Rachel Hart, Nicholas Serniak, Raphael Byrne, Burt Cagir

**Affiliations:** Department of Surgery, Robert Packer Hospital, Sayre, PA

## Abstract

Strictureplasty, introduced in 1978, has been suggested as an alternative to small bowel resection for cases of intestinal obstruction involving multiple segments, such as in Crohn’s disease. Despite research supporting the operation, little is known about outcomes after strictureplasty. Specifically, 30-day readmission and reoperation rates, which are widely recognized as surrogates for quality of surgical care have not been studied for this procedure. We sought to study the rate of, as well as factors associated with 30-day outcomes, including readmission and reoperation.

**METHODS:** We used the National Surgical Quality Improvement Program (NSQIP) participant user files (PUF) for the years 2012-2020. Primary Current Procedural Terminology (CPT) code “44615” was used to identify patients undergoing strictureplasty as their principal procedure. The outcomes of interest included related thirty-day readmissions and reoperations, non-routine discharge, and prolonged length of stay (LOS). Multivariable analyses were performed to identify factors associated with each outcome.

**RESULTS:** A total of 535 patients were identified with mean age 44.14 years (SD= 15.74). 52.5% were female. Thirty-day related readmission and reoperation rates were 9% (n=48) and 6% (n=32) respectively. Average LOS was 8.16 days (SD= 6.9). Non-routine discharge rate was4.2% (n=21). On multivariable logistic regression, factors associated with 30-day readmissions included longer operative time (OR 1.00, 95% CI 1.00-1.01, p=0.0008) and any surgical site infection (SSI) (OR 6.4, 95% CI 2.89-14.2, p<0.001). Increased LOS (OR 1.7, 95%CI 1.03-1.11, p=0.0009) and SSI (OR 28.1, 95%CI 10.44-75.47, p<0.001) were associated with 30-day related reoperation. Predictors of non-routine discharge included older age (65+ vs 18-40: OR 30.72, 95%CI 3.14-300.74, p=0.003) and longer LOS (OR 1.09, 95%CI 1.03-1.14, 0.0009). Factors associated with increased odds of prolonged LOS included higher ASA class (OR 3.29, 95%CI 2.10-5.14, p<0.001), longer operative time (1.00, 95%CI 1.00-1.01, p=0.0008) and SSI (OR 3.05, 95%CI 1.57-5.90, p=0.0009). Factors associated with lower odds of prolonged LOS included male sex (OR 0.56, 96%CI 0.36-0.88, p=0.011) and preoperative steroid use (OR 0.62, 95%CI 0.38-0.99, p=0.04).

**CONCLUSION:** Our analyses indicate that among patients undergoing strictureplasty, longer operative time, wound complications, and longer length of stay are associated with increased odds of readmission and reoperation within 30 days. These findings may be important for hospitals, providers, payors, and other stakeholders in further refining standards of quality of care for patients undergoing strictureplasty.

## INTRODUCTION

Small bowel resection has been suggested as an alternative to strictureplasty for the treatment of strictures related to Crohn’s disease (CD). Extensive resections with microscopically free margins have been proven to be unnecessary for CD as they do not reduce recurrence rates when compared to conservative resections. Strictureplasty leaves the diseased part of the intestine in place, but the need for additional surgery is unchanged.^1–6^

Strictureplasty, which Lee first proposed in 1978 as a way to treat obstructive CD while maintaining intestine length, has been established as a reliable and secure procedure.^7–9^ However, similar to other bowel-sparing surgeries, outcomes of strictureplasty have not been studied extensively. The literature is sparse for 30-day postoperative outcomes of this procedure in Crohn’s disease. We sought to query hospital and 30-day outcomes for strictureplasty for Crohn’s disease, using a national surgical quality registry Postoperative outcome, including readmissions and reoperations, have been used by National Quality Forum as surrogates for quality of care and are thus important for providers, hospitals and other stakeholders, as they help improve hospital discharge policies and post-operative care.

## METHODS

### Data Source

The National Surgical Quality Improvement Project (NSQIP) participant user files (PUF) for the years 2012-2020 were queried. We excluded prior years because variables pertaining to reasons for readmission and reoperation (including diagnosis codes at the time of readmission and reoperation) were first included in 2012.

### Cohort

Patients undergoing strictureplasty were first queried using the CPT code “44615” in either the main CPT column, or the 1st three columns in “Concurrent CPT” set and the “Other CPT” columns. Next, among patients undergoing strictureplasty, we only selected those with diagnosis codes for intestinal obstruction due to Crohn’s disease using International Classification of Disease (ICD) Clinical-Modification (CM) version 9 and 10. There were not any specific ICD-9 codes related to intestinal obstruction or structures secondary to Crohn’s disease. Therefore, we used the same set of ICD-9 diagnosis codes (550.x-558.x) as a previous study.^9^ For ICD-10, we used the diagnosis codes (K50-K50.x).

### Covariates of Interest

Variables of interest included demographic variables including age, sex, BMI (derived using weight and height), race; presence of comorbidities including diabetes, hypertension, dyspnea, COPD, smoking, ascites, functional status, weight loss, bleeding disorder, hypoalbuminemia and anemia; operative and Crohn’s related variables including steroid use (defined as patients requiring regular administration of oral or parenteral corticosteroid medications (e.g., Prednisone, Decadron) in the 30 days prior to surgery for a chronic medical condition (e.g., COPD, asthma, rheumatologic disease, rheumatoid arthritis, inflammatory bowel disease), American Society of Anesthesiology (ASA) classification, transfer status and elective or emergent case status.^10^

### Outcomes of Interest

Outcomes of interest were hospital data including prolonged length of stay (LOS), non-routine discharge, and 30-day outcomes including related readmission and reoperation. Prolonged LOS was defined as length of stay greater than the 75th percentile. We first calculated the median length of stay along with interquartile range, and subsequently coded all LOS greater than the 75th percentile (or 3rd quartile) as prolonged LOS. Non-routine discharge was defined as any discharge other than “home routine” or “a facility which was home”. Patients who died during the hospital stay were excluded from this analysis.

For related readmission and reoperation, we used the variable READMRELATED1 and RETORRELATED, respectively. READMRELATED1 is positive when a patient has a readmission related to their previous procedure, while RETORRELATED is positive for patients who have a return to the OR related to their previous procedure. Only the first readmission and reoperation were analyzed. We also queried reasons for reoperation using the ICD code assigned at the time of reoperation and classified them appropriately.

## RESULTS

A total of 535 patients were identified undergoing strictureplasty for Crohn’s disease between 2012 and 2020. The mean age of the cohort was 44.14 years (SD= 15.74) and 52.5% were females. The mean BMI was 24.41 (SD=6.31). A majority were White Caucasian patients (85%, n=455), followed by African Americans (5.6%, n=30) and Asians (1.5%, n=8). Hypertension was present in 16.3% (n=87), history of smoking in 13.6% (n=73), history of weight loss in 8.8% (n=47) and a bleeding disorder in 2.4% (n=12). Other comorbidities were less frequently present. Almost two-thirds of the cohort were on steroids (63.2%, n=338) preoperatively. Most patients were ASA 2 (55.1%, n=294). Of the remaining patients, 41.8% (n=223) were ASA 3 (Severe disturbance) and 2.1% (n=11) were ASA 4, and 1.1% (n=6) were ASA 1. Most patients (77.2%, n=413) had an elective procedure, while 22.6% (n=121) had an emergent procedure. Nearly half of the cohort (49.8%, n=202) had normal preoperative albumin levels; however, 29.1% (n=118) and 21.2% (n=86) had modest and severe hypoalbuminemia, respectively. The albumin level was missing for 129 patients (add %) and was therefore not included as a covariate in our multivariable analysis. 32.5% (n=163) and 8% (n=40) of patients had mild and moderate/severe anemia, respectively. These results are summarized in **Table 1**.

**Table 1:** Demographic Characteristics and Comorbidities of the Cohort

|  |  |
| --- | --- |
|  | Overall (N=535) |
| **Age** |  |
| Mean (SD) | 44.138 (15.736) |
| Range | 18 - 90 |
| **Age Groups** |  |
| 18-40 | 260 (48.6%) |
| 41-65 | 220 (41.1%) |
| 65+ | 55 (10.3%) |
| **Sex** |  |
| Female | 254 (47.5%) |
| Male | 281 (52.5%) |
| **BMI** |  |
| N-Miss | 4 |
| Mean (SD) | 24.408 (6.309) |
| Range | 13.468 - 70.105 |
| **Diabetes** | 21 (3.9%) |
| **Race** |  |
| American Indian or Alaska Native | 1 (0.2%) |
| Asian | 8 (1.5%) |
| Black or African American | 30 (5.6%) |
| White | 455 (85.0%) |
| Some Other Race | 1 (0.2%) |
| Unknown/Not Reported | 40 (7.5%) |
| **Hypertension** | 87 (16.3%) |
| **Dyspnea** | 6 (1.1%) |
| **Smoker** | 73 (13.6%) |
| **Hx of COPD** | 5 (0.9%) |
| **Ascites** | 3 (0.6%) |
| **Functional Status** |  |
| Independent | 533 (99.6%) |
| Partially Dependent | 2 (0.4%) |
| **Weight Loss** | 47 (8.8%) |
| **Bleeding Disorder** | 13 (2.4%) |
| **On Steroid** | 338 (63.2%) |
| **ASA Classification** |  |
| N-Miss | 1 |
| 1-No Disturb | 6 (1.1%) |
| 2-Mild Disturb | 294 (55.1%) |
| 3-Severe Disturb | 223 (41.8%) |
| 4-Life Threat | 11 (2.1%) |
| **Transfer Status** |  |
| From acute care hospital inpatient | 18 (3.4%) |
| Not transferred (admitted from home) | 511 (95.5%) |
| Nursing home - Chronic care - Intermediate care | 1 (0.2%) |
| Outside emergency department | 5 (0.9%) |
| **Elective or Emergent Surgery** |  |
| Emergent | 121 (22.6%) |
| Elective | 413 (77.2%) |
| Unknown | 1 (0.2%) |
| **Hypo-Albuminemia** |  |
| N-Miss | 129 |
| Normal | 202 (49.8%) |
| Modest hypoalbuminemia | 118 (29.1%) |
| Severe hypoalbuminemia | 86 (21.2%) |
| **Anemia** |  |
| N-Miss | 34 |
| No Anemia | 298 (59.5%) |
| Mild Anemia | 163 (32.5%) |
| Moderate/Severe Anemia | 40 (8.0%) |

The mean operative time was 195.5 minutes (SD=93.3) Four patients (0.7%, n=4)) required a post-operative blood transfusion. Most patients had a clean/contaminated wound class (66.7%, n=357). 21.3% (n=114) had a contaminated wound, 9.9% (n=53) had a dirty/infected wound, and 2.1% (n=11) had a clean wound. The average length of stay was 8.19 days (SD=6.9). The third quartile (75th percentile) for LOS was 9.5 days. Therefore, LOS greater than 9.5 days was regarded as prolonged LOS. A total of 512 (95.7%) patients were discharged home. Two patients died and 3 patients did not have discharge information available. Eighteen patients (3.4%) had a non-routine discharge. Of these, four patients were discharged to a rehabilitation facility, three patients were discharged to a separate acute care facility and eleven patients were discharged to a skilled care facility. Twenty-two patients (4.1%) had a superficial surgical site infection (SSI), 9 patients (1.7%) had a deep SSI, and 31 (5.8%) had an organ/space infection. Six patients (1.1%) had sepsis within 30 days. Thirty-day related readmission and reoperation rates were 9% (n=48) and 6% (n=32) respectively. These results are summarized in **Table 2**. On multivariable logistic regression, factors associated with 30-day readmissions included longer operative time (OR 1.00, 95% CI 1.00-1.01, p=0.0008) and surgical site infection (SSI) (OR 6.4, 95% CI 2.89-14.2,p<0.001) (**Figure 1, Table 3)**, while longer LOS (OR 1.7, 95%CI 1.03-1.11, p=0.0009) and SSI (OR 28.1, 95%CI 10.44-75.47, p<0.00) were associated with related reoperation (**Figure 2, Table 3)**. Predictors of non-routine discharge included older age (65+ vs 18-40: OR 30.72, 95%CI 3.14-300.74, p=0.003) and longer LOS (OR 1.09, 95%CI 1.03-1.14, 0.0009) (**Figure 3, Table 4)**.Predictors of prolonged LOS included higher ASA class (OR 3.29, 95%CI 2.10-5.14, p<0.001), longer operative time (1.00, 95%CI 1.00-1.01, p=0.0008) and SSI (OR 3.05, 95%CI 1.57-5.90, p=0.0009). Predictors of decreased LOS included male sex (OR 0.56, 96%CI 0.36-0.88, p=0.011) and steroid use (OR 0.62, 95%CI 0.38-0.99, p=0.04) (**Figure 4, Table 4)**.

**Table 2:**
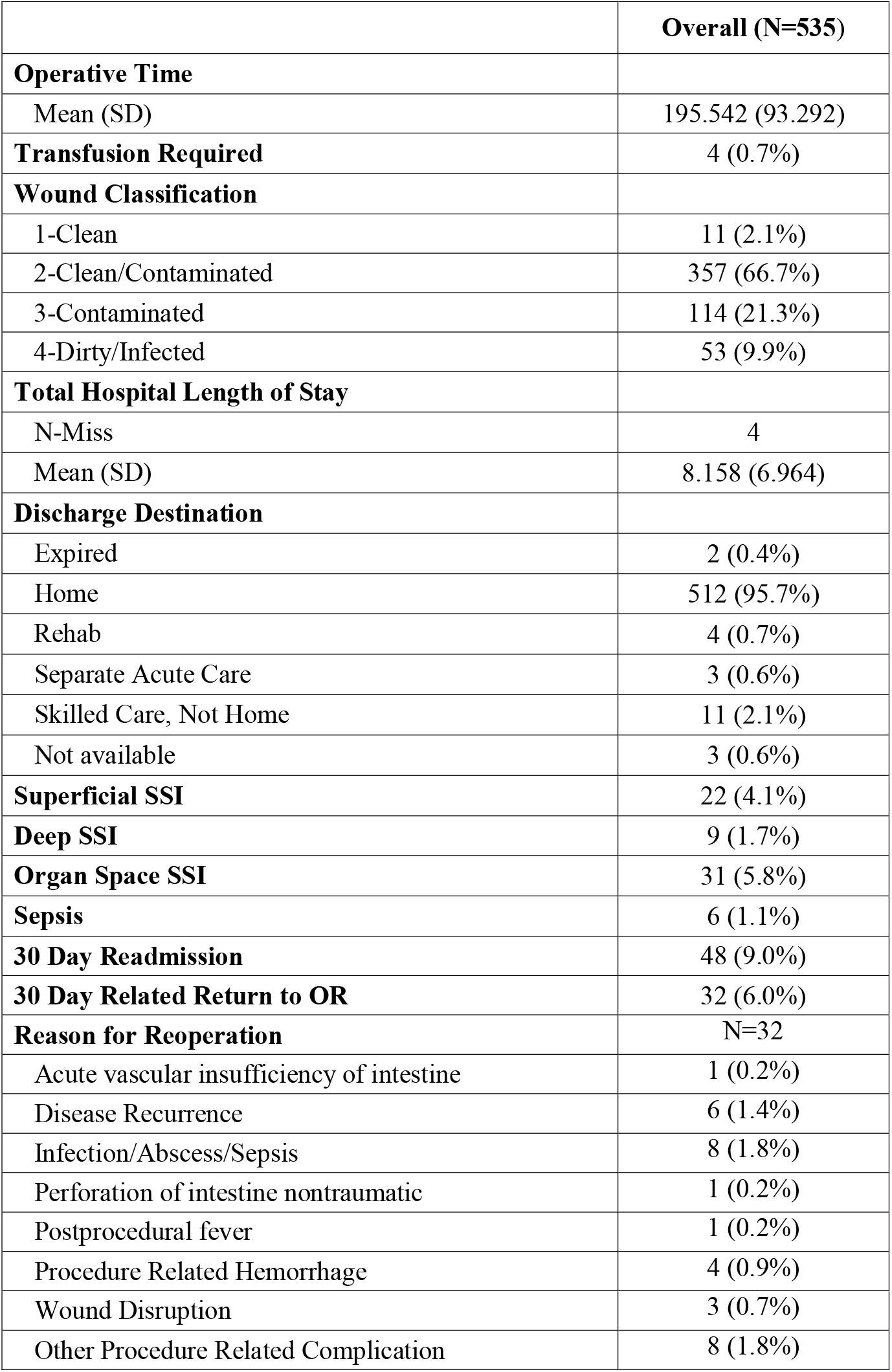
Summary of In-Hospital and 30-Day Outcomes after Strictureplasty

**Figure 1:**
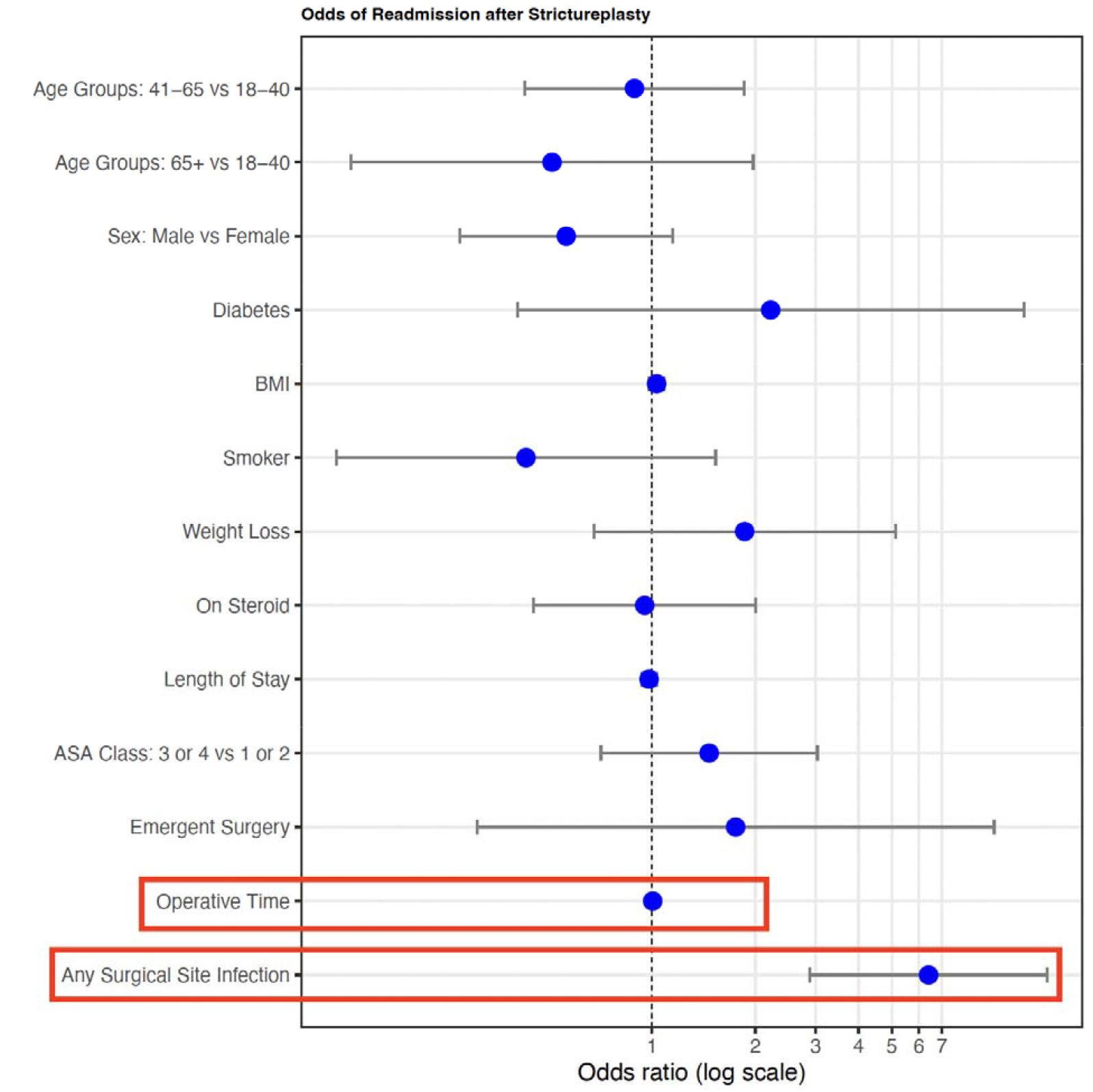
Multivariable Logistic Regression Model for 30-Day Related Readmission

**Table 3:** Multivariable Logistic Regression Analysis 30-Day Related Reoperation and Readmissions

| Covariate | 30-Day Related Reoperation |  |  |  | 30-Day Related Readmission |  |  |  |
| --- | --- | --- | --- | --- | --- | --- | --- | --- |
|  | Odds Ratio | Lower 95% CI | Upper 95% CI | p-value | Odds Ratio | Lower 95% CI | Upper 95% CI | p-value |
| Age Groups: 41-65 vs 18-40 | 0.50 | 0.18 | 1.39 | 0.1842 | 0.89 | 0.43 | 1.86 | 0.7535 |
| Age Groups: 65+ vs 18-40 | 0.49 | 0.11 | 2.18 | 0.3502 | 0.51 | 0.13 | 1.97 | 0.3299 |
| Sex: Male vs Female | 1.95 | 0.77 | 4.98 | 0.1616 | 0.56 | 0.28 | 1.15 | 0.1133 |
| Diabetes | 3.61 | 0.46 | 28.18 | 0.2214 | 2.22 | 0.40 | 12.14 | 0.3587 |
| BMI | 1.03 | 0.96 | 1.11 | 0.3551 | 1.03 | 0.98 | 1.08 | 0.2243 |
| Smoker | 0.71 | 0.17 | 2.93 | 0.6358 | 0.43 | 0.12 | 1.53 | 0.1926 |
| Weight Loss | 1.94 | 0.54 | 6.99 | 0.3118 | 1.86 | 0.68 | 5.12 | 0.2289 |
| On Preoperative Steroid | 1.48 | 0.53 | 4.10 | 0.4551 | 0.95 | 0.45 | 2.01 | 0.8966 |
| Length of Stay | 1.07 | 1.03 | 1.11 | **0.0009** | 0.98 | 0.93 | 1.03 | 0.4275 |
| ASA Class: 3 or 4 vs 1 or 2 | 0.90 | 0.35 | 2.33 | 0.8255 | 1.47 | 0.71 | 3.03 | 0.3008 |
| Emergent Surgery | 1.55 | 0.23 | 10.64 | 0.6536 | 1.75 | 0.31 | 9.94 | 0.5261 |
| Operative Time | 0.99 | 0.99 | 1.00 | 0.050 | 1.00 | 1.00 | 1.01 | **0.0101** |
| Any Surgical Site Infection | 28.08 | 10.44 | 75.47 | **<0.0001** | 6.40 | 2.89 | 14.20 | **<0.0001** |

**Figure 2:**
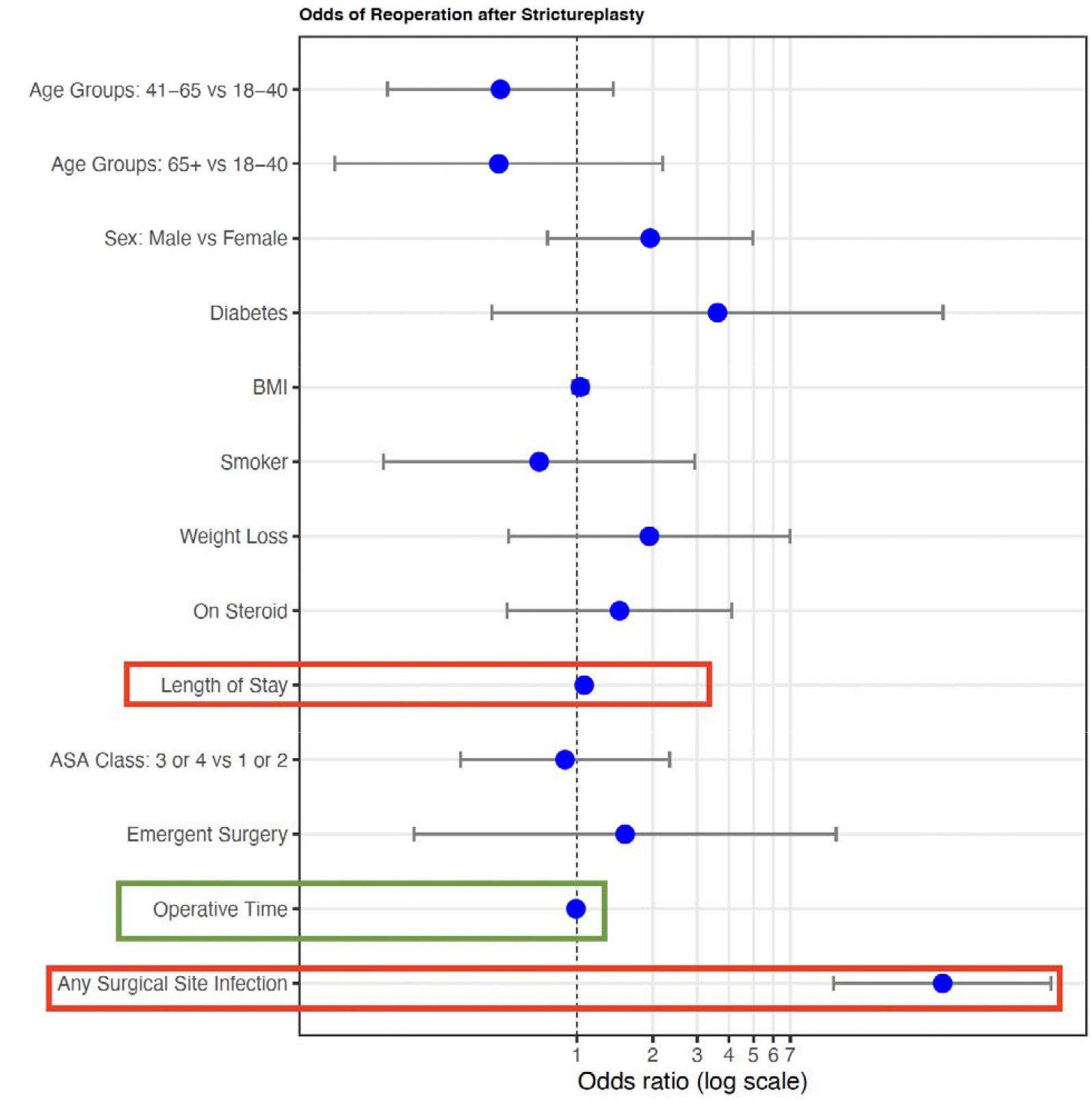
Multivariable Logistic Regression Model for 30-Day Related Reoperation

**Figure 3:**
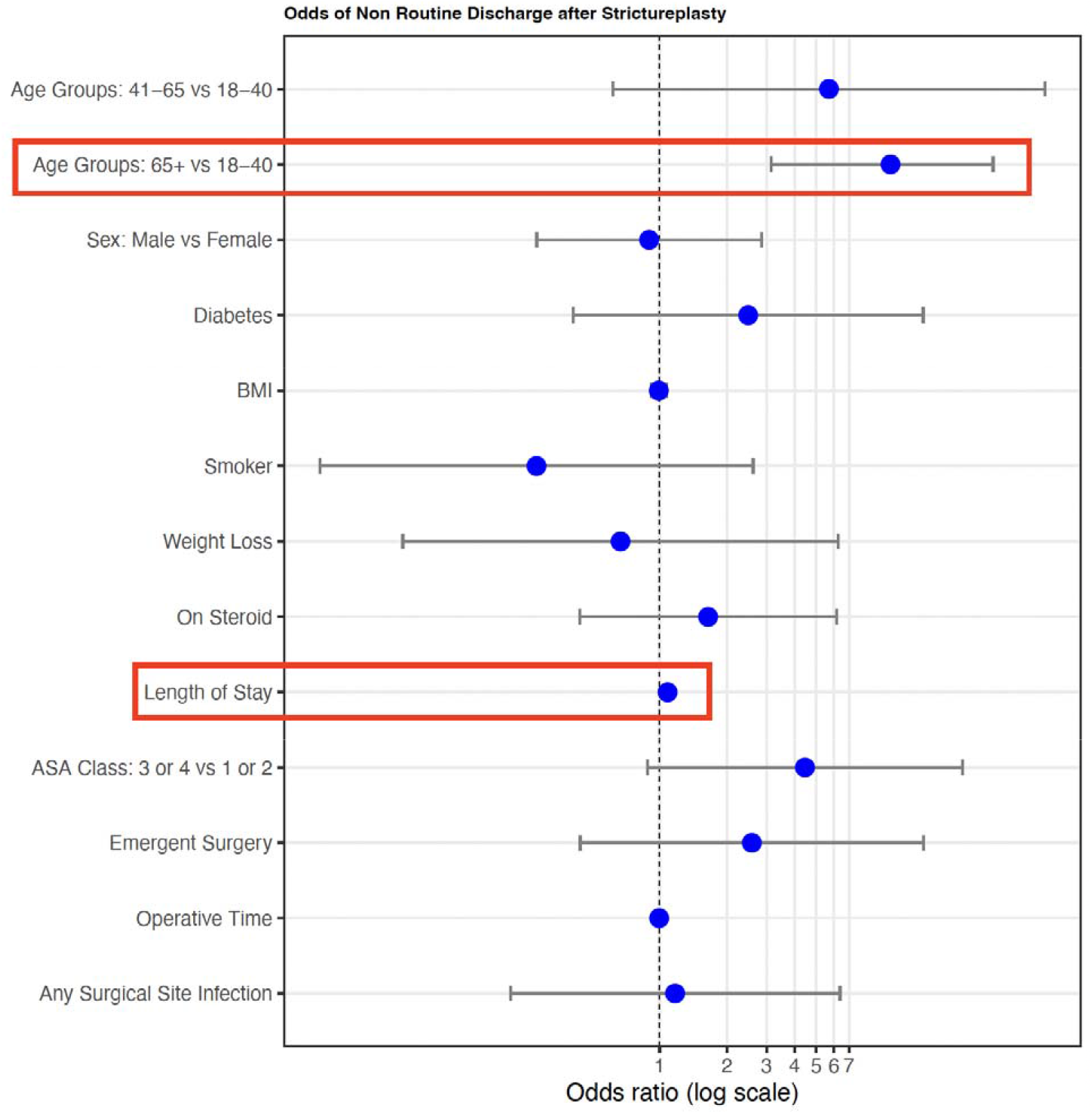
Multivariable Logistic Regression Model for Non-Routine Discharge

**Table 4:** Multivariable Logistic Regression Analysis for Non-Routine Discharge and Prolonged Length of Stay

| Covariate | Non-Routine Discharge |  |  |  |  | Prolonged Length of Stay |  |  |  |
| --- | --- | --- | --- | --- | --- | --- | --- | --- | --- |
|  | Odds Ratio | Lower 95% CI | Upper 95% CI | p-value |  | Odds Ratio | Lower 95% CI | Upper 95% CI | p-value |
| Age Groups: 41-65 vs 18-40 | 5.70 | 0.62 | 52.47 | 0.1247 |  | 0.93 | 0.58 | 1.50 | 0.7762 |
| Age Groups: 65+ vs 18-40 | 30.72 | 3.14 | 300.74 | **0.0033** |  | 1.36 | 0.64 | 2.89 | 0.4202 |
| Sex: Male vs Female | 0.90 | 0.28 | 2.85 | 0.8547 |  | 0.56 | 0.36 | 0.88 | **0.0115** |
| Diabetes | 2.48 | 0.41 | 15.01 | 0.3214 |  | 2.10 | 0.58 | 7.55 | 0.2561 |
| BMI | 0.99 | 0.92 | 1.07 | 0.8391 |  | 0.95 | 0.91 | 1.00 | **0.046** |
| Smoker | 0.28 | 0.03 | 2.61 | 0.2652 |  | 1.54 | 0.85 | 2.78 | 0.1552 |
| Weight Loss | 0.67 | 0.07 | 6.27 | 0.7249 |  | 1.34 | 0.65 | 2.79 | 0.4286 |
| On Preoperative Steroid | 1.65 | 0.44 | 6.16 | 0.4592 |  | 0.62 | 0.38 | 0.99 | **0.0453** |
| Length of Stay | 1.09 | 1.03 | 1.14 | **0.0009** |  | NA | NA | NA | NA |
| ASA Class: 3 or 4 vs 1 or 2 | 4.45 | 0.88 | 22.42 | 0.0701 |  | 3.29 | 2.10 | 5.14 | **<0.0001** |
| Emergent Surgery | 2.58 | 0.44 | 15.04 | 0.2921 |  | 1.65 | 0.54 | 5.06 | 0.3806 |
| Operative Time | 1.00 | 0.99 | 1.00 | 0.2034 |  | 1.00 | 1.00 | 1.01 | **0.0008** |
| Any Surgical Site Infection | 1.17 | 0.22 | 6.38 | 0.854 |  | 3.05 | 1.57 | 5.90 | **0.0009** |

**Figure 4:**
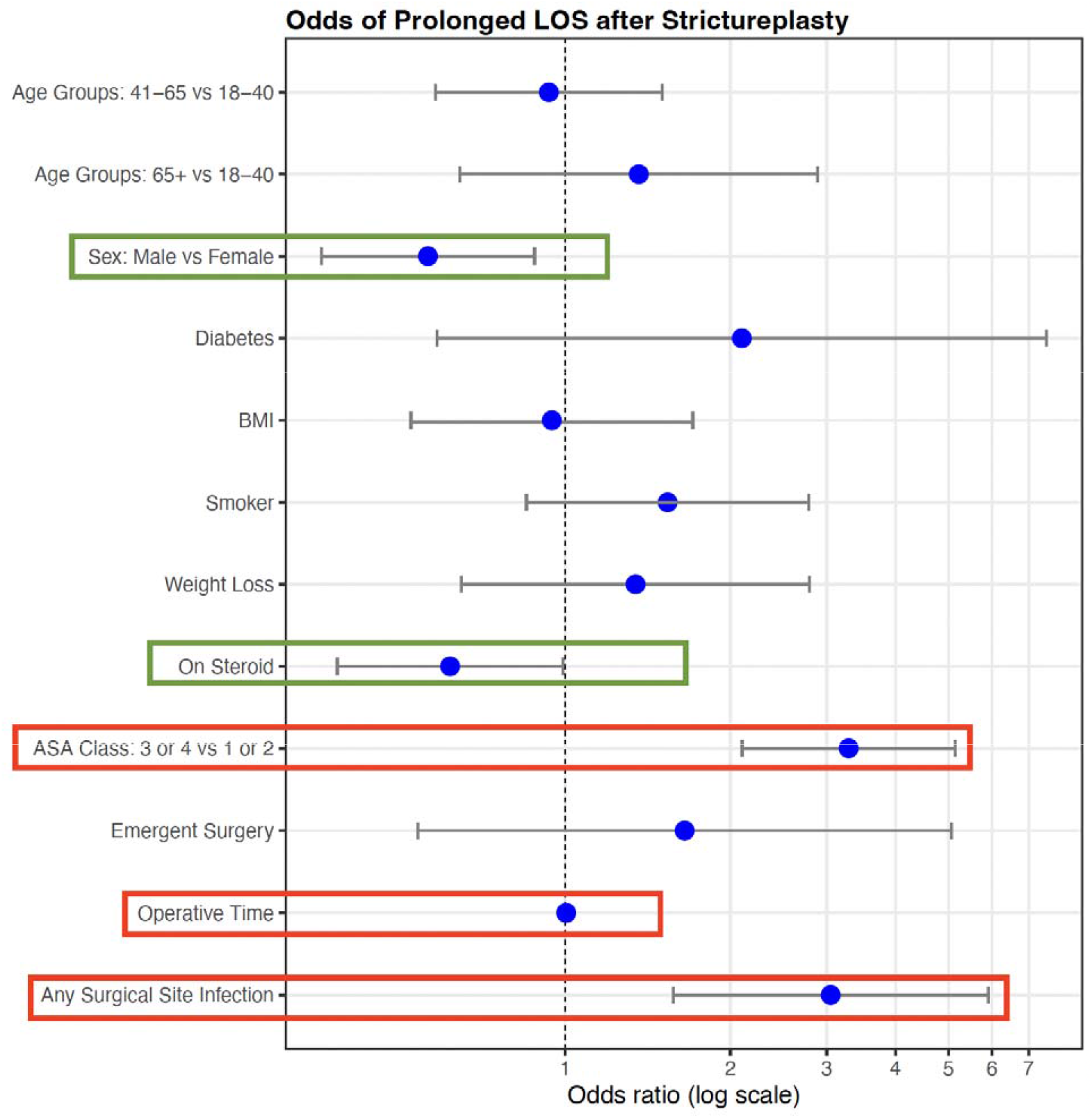
Multivariable Logistic Regression Model for Prolonged Length of Stay

## DISCUSSION

Here we present results from NSQIP data on strictureplasty for Crohn’s disease and factors associated with hospital and 30-day outcomes.

Hospital length of stay is an important operative quality metric. Physical recovery following surgery depends on several variables, including patient age, congenital disease, psychological and social well-being, type of operation, wound size, type of pain, and length of surgery. Recovery may be hampered if vital signs and circulation are not sufficiently stable during surgery. Effective postoperative care should therefore aim to shorten hospital stays and decrease complications.^11,12^ W investigated factors associated with prolonged length of stay after strictureplasty for Crohn’s disease. We found that higher ASA class, longer operative time, and incidence of SSI were associated with increased odds of prolonged length of stay, while male sex and preoperative steroid use reduced the odds. Higher ASA class reflects poor systemic status of the patient, including presence of comorbidities such as poorly controlled diabetes mellitus/hypertension, COPD and reduced ejection fraction, among others. These comorbidities may require stabilization pre- and post-operatively, contributing to increased stay in the hospital prior to discharge. Luong et al. presented results from a multicenter retrospective cohort of patients with CD undergoing abdominal operations in four tertiary centers in Denmark. They found age, preoperative abscess, and previous laparotomy/laparoscopy to be predictive of prolonged hospital stay. However, it is important to note that their definition of prolonged LOS was an LOS above median, while ours was based on a more traditional definition of LOS >75th percentile. We agree with Luong’s results that infection or abscess prolongs LOS. We also found male sex to be associated with lower odds of prolonged LOS. Previous studies have indicated females to be at higher risk for adverse outcomes after strictureplasty for Crohn’s, though the reasons remain unclear.^13^

Discharge disposition after major colorectal surgery has been well studied with recent improvements in multimodal perioperative care pathways namely ERAS (Enhanced Recovery After Surgery).^14–17^ Although postoperative morbidity is predicted to have an impact on the requirement for longer-term care, it is unclear from the literature if these predictions differ between patients with or without complications even with application of ERAS pathways. An additional health policy concern is the financial burden brought on by various post-acute care program variations.^18^ Our results indicate that older patients and prolonged length of stay may be associated with higher odds of adverse discharge. Our results are corroborated by previous studies,^19–21^ including work by Al-Mazrou et al. who queried patients undergoing colectomy and found older age and longer length of stay to predict non-home discharge.^18^ Older patients, including those on Medicare, may be required to stay in hospitals for longer before discharge to rehabilitation. Other factors contributing to prolonged hospital stay may include higher incidence of comorbidities in older patients. Moreover, recent advancements in enhanced recovery after surgery (ERAS) protocols have garnered much interest and thus, some of the variability in outcomes in our results may be attributable to the institution.

Longer operative time, incidence of SSI, and prolonged length of stay were associated with higher odds of readmission and reoperation within 30 days. Longer operative time is likely a surrogate for surgical complexity. The strongest indicator for readmission and reoperation was a surgical site infection. The rate of wound complication overall (9.7%) was comparable to previous results including a meta-analysis by Tichansky et al. who found an overall wound complication rate of 8.1% based on results from 506 patients undergoing 1,825 strictureplasties.^22^ It is important to note that Tichansky et al. also separately reported the rate of abscess in their meta-analysis to be 10.8%. Our overall rate was inclusive of “Organ/Space” infection which may include abscesses. However, if we only include superficial and deep surgical site infections, our rate falls to 5.8%. Improved antiseptic techniques may be responsible for this decrease.

Wound related complication was the most common reason for reoperation (2.5%, n=11). The second most common reason for a reoperation was disease recurrence (1.4%, n=6). According to previous studies with 3–5-year follow-up, the rate of stricture recurrence ranges from 23-32%, depending on the technique. However, it is interesting to note that up to 1.5% may present with the same complaint within 30 days. This may indicate inadequate resolution of stricture(s) in the primary surgery and warrants further investigation.

The role of preoperative steroids in patients with Crohn’s disease undergoing surgery has been controversial. Several studies have shown their use to be associated with increased risk of septic complication and anastomotic leakage,^23–27^ while others have shown no increase in risk. ^28–30^ Our results did not demonstrate an increase in risk of readmission or reoperation with preoperative steroid use. Of note, steroid use is marked positive in NSQIP when the patient received regular administration of oral or parenteral corticosteroid medications (e.g., Prednisone, Decadron) in the 30 days prior to surgery for a chronic medical condition (e.g., COPD, asthma, rheumatologic disease, rheumatoid arthritis, inflammatory bowel disease). In other literature on Crohn’s disease, preoperative steroid use is defined as prednisone doses (or its equivalent) between 5 and 40 mg per day, 14 to 60 days prior to surgery, for at least 4 weeks, but not longer than 3 months. ^24,29,31–33^ Therefore, the reported rate of steroid use in our study may be an underestimate based on these definitions.

## LIMITATIONS

This is one of the most comprehensive studies performed on 30-day outcomes after strictureplasty for Crohn’s disease using a national surgical quality registry. There has been one previous study using data from NSQIP,^9^ however, the authors did not perform multivariable analyses. Nevertheless, there are some limitations to our study. We relied on CPT codes for defining our cohort, however previous studies have shown coding errors to be common. Therefore, there may be some included patients who did not receive a strictureplasty. Moreover, we relied on diagnosis codes for defining the reason for reoperation. Finally, while we did report the rate of anemia and hypoalbuminemia in our results, we were unable to assess their impact on outcomes due to missing values. Therefore, there may be some residual confounding factors that may not have been accounted for in our multivariable model.

## CONCLUSION

Our analyses indicate that among patients undergoing strictureplasty, longer operative time – likely reflecting complex procedures wound complications, and longer length of stay increase odds of readmission and reoperation within 30 days. We also found that older age and prolonged hospital LOS may be predictive of adverse discharge, while higher ASA class, longer operative times, and SSI are associated with longer hospital stay. These findings may be important for hospitals, providers, payors, and other stakeholders in further refining standards of quality of care for patients undergoing strictureplasty.

## Data Availability

All data produced are available online at ACS website which is provided as NSQIP data set.

## REFERENCES

1. Satsangi. Satsangi J, Silverberg MS, Vermeire S, Colombel JF. The Montreal classification of inflammatory bowel disease: controversies, consensus, and …. Gut.

2. Louis, E. et al. Behaviour of Crohn’s disease according to the Vienna classification: changing pattern over the course of the disease. Gut 49, 777–782 (2001).

3. Solberg, I. C. et al. Clinical course in Crohn’s disease: results of a Norwegian population-based ten-year follow-up study. Clin. Gastroenterol. Hepatol. 5, 1430–1438 (2007).

4. Thia, K. T., Sandborn, W. J., Harmsen, W. S., Zinsmeister, A. R. & Loftus, E. V., Jr. Risk factors associated with progression to intestinal complications of Crohn’s disease in a population-based cohort. Gastroenterology 139, 1147–1155 (2010).

5. Rieder, F., Zimmermann, E. M., Remzi, F. H. & Sandborn, W. J. Crohn’s disease complicated by strictures: a systematic review. Gut 62, 1072–1084 (2013).

6. Peyrin-Biroulet, L., Loftus, E. V., Jr, Colombel, J.-F. & Sandborn, W. J. The Natural History of Adult Crohn’s Disease in Population-Based Cohorts. Official journal of the American College of Gastroenterology | ACG 105, 289 (2010).

7. Lee, E. C. & Papaioannou, N. Minimal surgery for chronic obstruction in patients with extensive or universal Crohn’s disease. Ann. R. Coll. Surg. Engl. 64, 229–233 (1982).

8. Yamamoto, T., Fazio, V. W. & Tekkis, P. P. Safety and efficacy of strictureplasty for Crohn’s disease: a systematic review and meta-analysis. Dis. Colon Rectum 50, 1968–1986 (2007).

9. Geltzeiler, C. B. et al. Strictureplasty for Treatment of Crohn’s Disease: an ACS-NSQIP Database Analysis. J. Gastrointest. Surg. 19, 905–910 (2015).

10. ACS NSQIP Participant Use Data File. ACS https://www.facs.org/quality-programs/data-and-registries/acs-nsqip/participant-use-data-file/.

11. Trang & Thosingha. Factors Associated with Recovery among Patients after Abdominal Surgery: ป้จจัย ที่ มี ผล ต่อ การ พื้นตัว ของ ผู้ ป่วย หลัง ผ่าตัด ช่อง ท้อง. Growth Perform. Yield Oyster Mushroom (Pleurotus Ostreatus) Differ. Substr. Compos. Buea South West Cameroon.

12. Udomkhwamsuk, W., Vuttanon, N. & Limpakan, S. Situational analysis on the recovery of patients who have undergone major abdominal surgery. Nurs Open 8, 140–146 (2021).

13. Dasari, B. V. M., Maxwell, R. & Gardiner, K. R. Assessment of complications following strictureplasty for small bowel Crohn’s Disease. Ir. J. Med. Sci. 179, 201–205 (2010).

14. Cook, C. H., Martin, L. C., Howard, B. & Flancbaum, L. J. Survival of critically ill surgical patients discharged to extended care facilities. J. Am. Coll. Surg. 189, 437–441 (1999).

15. Legner, V. J., Massarweh, N. N., Symons, R. G., McCormick, W. C. & Flum, D. R. The significance of discharge to skilled care after abdominopelvic surgery in older adults. Ann. Surg. 249, 250–255 (2009).

16. Marcantonio, E. R. et al. Outcomes of older people admitted to postacute facilities with delirium. J. Am. Geriatr. Soc. 53, 963–969 (2005).

17. Ottenbacher, K. J. et al. Thirty-day hospital readmission following discharge from postacute rehabilitation in fee-for-service Medicare patients. JAMA 311, 604–614 (2014).

18. Al-Mazrou, A. M., Chiuzan, C. & Kiran, R. P. Factors influencing discharge disposition after colectomy. Surg. Endosc. 32, 3032–3040 (2018).

19. Mohanty, S. et al. Risk of discharge to postacute care: a patient-centered outcome for the american college of surgeons national surgical quality improvement program surgical risk calculator. JAMA Surg. 150, 480–484 (2015).

20. Sacks, G. D. et al. Which Patients Require More Care after Hospital Discharge? An Analysis of Post-Acute Care Use among Elderly Patients Undergoing Elective Surgery. J. Am. Coll. Surg. 220, 1113–1121.e2 (2015).

21. Legner, V. J., Doerner, D., Reilly, D. F. & McCormick, W. C. Risk factors for nursing home placement following major nonemergent surgery. Am. J. Med. 117, 82–86 (2004).

22. Tichansky, D., Cagir, B., Yoo, E., Marcus, S. M. & Fry, R. D. Strictureplasty for Crohn’s disease. Dis. Colon Rectum 43, 911–919 (2000).

23. Sharma, A. & Chinn, B. T. Preoperative optimization of crohn disease. Clin. Colon Rectal Surg. 26, 75–79 (2013).

24. Alves, A. et al. Risk factors for intra-abdominal septic complications after a first ileocecal resection for Crohn’s disease: a multivariate analysis in 161 consecutive patients. Dis. Colon Rectum 50, 331–336 (2007).

25. Yamamoto, T., Allan, R. N. & Keighley, M. R. Risk factors for intra-abdominal sepsis after surgery in Crohn’s disease. Dis. Colon Rectum 43, 1141–1145 (2000).

26. Post, S. et al. Risks of intestinal anastomoses in Crohn’s disease. Ann. Surg. 213, 37–42 (1991).

27. Aberra, F. N. et al. Corticosteroids and immunomodulators: postoperative infectious complication risk in inflammatory bowel disease patients. Gastroenterology 125, 320–327 (2003).

28. Bruewer, M. et al. Preoperative steroid administration: effect on morbidity among patients undergoing intestinal bowel resection for Crohńs disease. World J. Surg. 27, 1306–1310 (2003).

29. Canedo, J. et al. Surgical resection in Crohn’s disease: is immunosuppressive medication associated with higher postoperative infection rates? Colorectal Dis. 13, 1294–1298 (2011).

30. Mascarenhas, Nunoo & Asgeirsson. Outcomes of ileocolic resection and right hemicolectomies for Crohn’s patients in comparison with non-Crohn’s patients and the impact of perioperative …. journal of surgery.

31. Subramanian, V., Saxena, S., Kang, J.-Y. & Pollok, R. C. G. Preoperative steroid use and risk of postoperative complications in patients with inflammatory bowel disease undergoing abdominal surgery. Am. J. Gastroenterol. 103, 2373–2381 (2008).

32. Tzivanakis, A. et al. Influence of risk factors on the safety of ileocolic anastomosis in Crohn’s disease surgery. Dis. Colon Rectum 55, 558–562 (2012).

33. Mascarenhas, Nunoo & Asgeirsson. … resection and right hemicolectomies for Crohn’s patients in comparison with non-Crohn’s patients and the impact of perioperative immunosuppressive therapy …. journal of surgery.

